# Reconstructing transmission chains of SARS-CoV-2 amid multiple outbreaks in a geriatric acute-care hospital

**DOI:** 10.1101/2022.01.07.22268729

**Authors:** Mohamed Abbas, Anne Cori, Samuel Cordey, Florian Laubscher, Tomás Robalo Nunes, Ashleigh Myall, Julien Salamun, Philippe Huber, Dina Zekry, Virginie Prendki, Anne Iten, Laure Vieux, Valérie Sauvan, Christophe E. Graf, Stephan Harbarth

## Abstract

**Background:** There is ongoing uncertainty regarding transmission chains and the respective roles of healthcare workers (HCWs) and elderly patients in nosocomial outbreaks of severe acute respiratory syndrome coronavirus 2 (SARS-CoV-2) in geriatric settings.

**Methods:** We performed a retrospective cohort study including patients with nosocomial coronavirus disease 2019 (COVID-19) in four outbreak-affected wards, and all SARS-CoV-2 RT-PCR positive HCWs from a Swiss university-affiliated geriatric acute-care hospital that admitted both Covid-19 and non-Covid-19 patients during the first pandemic wave in Spring 2020. We combined epidemiological and genetic sequencing data using a Bayesian modelling framework, and reconstructed transmission dynamics of SARS-CoV-2 involving patients and HCWs, in order to determine who infected whom. We evaluated general transmission patterns according to type of case (HCWs working in dedicated Covid-19 cohorting wards: HCW_covid_; HCWs working in non-Covid-19 wards where outbreaks occurred: HCW_outbreak_; patients with nosocomial Covid-19: patient_noso_) by deriving the proportion of infections attributed to each type of case across all posterior trees and comparing them to random expectations.

**Results:** During the study period (March 1 to May 7, 2020) we included 180 SARS-CoV-2 positive cases: 127 HCWs (91 HCW_covid_, 36 HCW_outbreak_) and 53 patients. The attack rates ranged from 10-19% for patients, and 21% for HCWs. We estimated that there were 16 importation events (3 patients, 13 HCWs) that jointly led to 16 secondary cases. Most patient-to-patient transmission events involved patients having shared a ward (97.6%, 95% credible interval [CrI] 90.4-100%), in contrast to those having shared a room (44.4%, 95%CrI 27.8-62.5%). Transmission events tended to cluster by type of case: patient_noso_ were almost twice as likely to be infected by other patient_noso_ than expected (observed:expected ratio 1.91, 95%CrI 1.08 – 4.00, **p** = 0.02); similarly, HCW_outbreak_ were more than twice as likely to be infected by other HCW_outbreak_ than expected (2.25, 95%CrI 1.00-8.00, **p** = 0.04). The proportion of infectors of HCW_covid_ were as expected as random. The proportions of high transmitters (≥2 secondary cases) were significantly higher among HCW_outbreak_ than patient_noso_ in the late phases (26.2% vs. 13.4%, p<2.2e-16) of the outbreak.

**Conclusions:** Most importation events were linked to HCW. Unexpectedly, transmission between HCW_covid_ was more limited than transmission between patients and HCW_outbreak_. This highlights gaps in infection control and suggests possible areas of improvements to limit the extent of nosocomial transmission.

## Introduction

Nosocomial acquisition of severe acute respiratory syndrome coronavirus 2 (SARS-CoV-2) in geriatric institutions and long-term care facilities (LTCFs) may account for large proportions of all declared coronavirus disease 2019 (Covid-19) cases in many countries, and contribute to a large extent to morbidity and mortality [1-4]. The reservoir of SARS-CoV-2 in healthcare environments may contribute to amplifying the pandemic [5], and as such, it is important to understand transmission dynamics in these settings.

The terms healthcare-associated, hospital-onset, and nosocomial Covid-19 reflect the uncertainty around defining and distinguishing community-versus healthcare-acquired Covid-19 cases [6]. Nevertheless, in some settings such as LTCFs and nursing homes these definitions are relatively straightforward. In other settings, such as those with a high turnover of patients, or where patients are admitted from the community and both Covid-19 and non-Covid-19 cases are hospitalised in the same institution, defining, and more importantly detecting cases is crucial to avoid cross-contamination. Determining sources and transmission pathways of infection may thus help improve infection prevention and control (IPC) strategies.

The role of healthcare workers (HCWs) in nosocomial Covid-19 is complex, as they can be victims and/or vectors of SARS-CoV-2 infection, and can acquire from or transmit to their peers and patients as well as the community [7, 8]. There is ongoing controversy and uncertainty surrounding the role of HCWs in infecting patients in nosocomial outbreaks, and in particular findings from acute-care hospitals cannot be applied directly to LTCFs and geriatric hospitals [9-12].

The aim of this study was to reconstruct transmission dynamics in several nosocomial outbreaks of SARS-CoV-2 involving patients and HCWs in a Swiss university-affiliated geriatric hospital that admitted both Covid-19 and non-Covid-19 patients during the first pandemic wave in Spring 2020.

## Methods

We performed a retrospective cohort study including patients with nosocomial COVID-19 in four outbreak-affected wards, as well as all SARS-CoV-2 RT-PCR positive HCWs from March 1 to May 7, 2020.

### Setting

The Hospital of Geriatrics, part of the Geneva University Hospitals (HUG) consortium, is a facility with 196 acute-care and 100 rehabilitation beds. During the first pandemic wave, a maximum of 176 acute-care beds were dedicated to admitting geriatric patients with Covid-19 who were not eligible for escalation of therapy (e.g. intensive care unit admission) [13]. During the same period, patients were also admitted for non-Covid-19 hospitalisations, and the rehabilitation beds were also open to convalescent Covid-19 patients. The implemented IPC measures are detailed in the Supplement. RT-PCR screening of SARS-CoV-2 on admission for all patients, even if asymptomatic or without clinical suspicion of Covid-19, started on April 01, 2020. From April 07, 2020, weekly screening surveys were performed in non-Covid wards, until May 30, 2020. HCWs from outbreak wards were encouraged to undergo PCR testing on nasopharyngeal swabs, even if asymptomatic between April 09 and April 16, 2020.

### Definitions

Healthcare-associated (HA) Covid-19 was defined by an onset of symptoms ≥ 5 days after admission in conjunction with a strong suspicion of healthcare transmission, in accordance with Swissnoso guidelines [14]. Patients with HA-Covid-19 were classified as “patient_noso_”, the others were assumed community-acquired or “patient_community_”. An outbreak was declared when there were ≥3 cases of HA-Covid-19 cases (HCWs and patients) with a possible temporal-spatial link [14]. HCWs were included in the outbreak investigation if they presented a positive RT-PCR for SARS-CoV-2. HCWs were classified as “HCW_covid_” if they worked in a Covid-19 cohorting ward admitting community-acquired cases, or “HCW_outbreak_” if they worked in a “non-Covid” ward (i.e. not admitting community-acquired Covid-19 cases) in which nosocomial outbreaks occurred.

### Data sources

The data used for this study were from the same sources as described previously [9]. First, we used prospectively collected data from the Swiss Federal Office of Public Health-mandated surveillance of hospitalised Covid-19 patients [1]. We also used prospectively collected data from HUG’s Department of Occupational Health for symptom-onset data as well as the Department of Human Resources (HR) for HCW shifts.

### Descriptive epidemiology

We produced an epidemic curve using dates of symptom onset; where these were unavailable (e.g. lack of symptoms), we imputed them with the median difference between date of symptom onset and date of nasopharyngeal swab.

### Microbiological methods

All COVID-19 cases in the outbreak were confirmed by RT-PCR on nasopharyngeal swabs. We performed SARS-CoV-2 whole genome sequencing (WGS) using an amplicon-based sequencing method in order to produce RNA sequences, as previously described [9] and summarised in the Supplement.

### Phylogenetic analysis

Sequence alignment was performed with MUSCLE (v3.8.31). The evolutionary analyses were conducted in MEGA X [15] using the Maximum Likelihood method and Tamura 3-parameter model [16]. All SARS-CoV-2 complete genomes sequenced by the Laboratory of Virology (HUG) in the context of city-wide epidemiological surveillance were integrated to the phylogenetic analysis in addition to the case samples of this outbreak analysis.

### Statistical analysis

We performed descriptive statistics with medians and interquartile ranges (IQR), and counts and proportions, as appropriate.

### Reconstruction of transmission trees

We combined epidemiological and genomic data to reconstruct who infected whom using the R package `outbreaker2` [17, 18], as described elsewhere [9] and in the Supplement. Briefly, the model uses a Bayesian framework, combining information on the generation time (time between infections in an infector/infectee pair), contact patterns, with a model of sequence evolution to probabilistically reconstruct the transmission tree (see Supplement).

Because formal contact tracing was limited during the study period, we constructed contact networks based on ward or room presence for patients based on their trajectories, and on HCW shifts obtained from HR. A contact was defined as simultaneous presence on the same ward on a given day (see Supplement). The manner by which `outbreaker2` handles these contacts is conservative in that it allows for A) non-infectious contacts to occur, and B) incomplete reporting of infectious contacts; the proportions of these are estimated in the model output.

Using the reconstructed transmission trees, we calculated the number of secondary (i.e. onwards) infections for each case, i.e. the individual reproductive number (*R*), which we stratified by epidemic phase (early or late with a cut off on April 09, 2020) and type of case (HCW_covid_, HCW_outbreak_, patient_noso_, patient_community_, see Supplement).

We assessed the role of each type of case in transmission by estimating the proportion of infections attributed to the type of case (*f*_*case*_), which we compared to the random expectation considering the prevalence of each type among cases (see Supplement). To better understand the transmission pathways between and within wards, we also estimated (for outbreak and non-outbreak wards), the proportion of infections attributed to infectors in the same ward. We also constructed a matrix representing ward-to-ward transmission. Patient movements between wards were constructed using the implementation of the `vistime` package (visualisation tool) as in the publication by Meredith et al. [19]. Statistical analyses were performed in R software version 4.0.3 (https://www.R-project.org/).

### Ethical considerations

The Ethics Committee of the Canton of Geneva (CCER), Switzerland, approved this study (CCER no. 2020-01330 and CCER no. 2020-00827).

## Results

During the study period, we included a total of 180 SARS-CoV-2 positive cases: 127 HCWs of whom 91 HCW_covid_, and 36 HCW_outbreak_, and 53 patients from the 4 outbreak wards. Of the 53 included patients, **post-hoc** epidemiological analysis showed that 4 of these were highly likely community-acquired Covid-19 (CA-Covid-19).

The remaining 49 nosocomial cases represented 20.2% (49/242) of all hospitalised COVID-19 patients, and 81.7% (49/60) of nosocomial Covid-19 cases of the Geriatric Hospital. The ward-level attack rates ranged from 10 to 19% among patients. Moreover, 21% of all HCWs in the geriatric hospital had a PCR-positive test. The epidemic curve is shown in Figure 1, and ward-level epidemic curves in Supplementary Figure S1. Characteristics of patients and HCWs are summarised in Tables 1 and 2, respectively.

**Table 1.** Characteristics of CoVID-19 patients with nosocomial acquisition

| Characteristics | All patients<br>(N = 49) |
| --- | --- |
| Female, n (%) | 28 (57.1) |
| Age, median (IQR) | 85.4 (83.5 – 89.3) |
| Asymptomatic, n (%) | 3 (6.1) |
| Onset of symptoms before swab date, n (%) | 12 (24.5) |
| Days from onset of symptoms to swab, median (IQR) | 0 (0-0) |
| Days from onset of symptoms to swab, mean (SD) | -0.29 (2.19) |

**Table 2.** Characteristics of SARS-CoV-2 RT-PCR positive HCWs

| Characteristics | All HCWs<br>(N = 127) |
| --- | --- |
| Female, n (%) | 92 (72.4) |
| Age, median (IQR) | 32.0 (43.3 – 54.8) |
| Profession, n (%) |  |
| Nurse | 57 (44.9) |
| Nurse assistant | 39 (30.7) |
| Doctor | 19 (15.0) |
| Care assistant | 4 (3.2) |
| Transporter | 4 (3.2) |
| Physical therapist | 2 (1.6) |
| Logopedist | 1 (0.8) |
| Medical student | 1 (0.8) |
| Asymptomatic, n (%) | 5 (3.9) |
| *missing data for 5* |  |
| Days from onset of symptoms to swab, median (IQR) | 1 (-2 to 21) |
| HCWs in Covid-19 wards | 1 (1-2) |
| HCWs in non-Covid (outbreak) wards | 1 (0-3) |
| Days from onset of symptoms to swab, mean (SD) | 1.91 (2.86) |
| HCWs in Covid-19 wards | 1.60 (1.78) |
| HCWs in non-Covid (outbreak) wards | 2.88 (4.84) |

**Figure 1.**
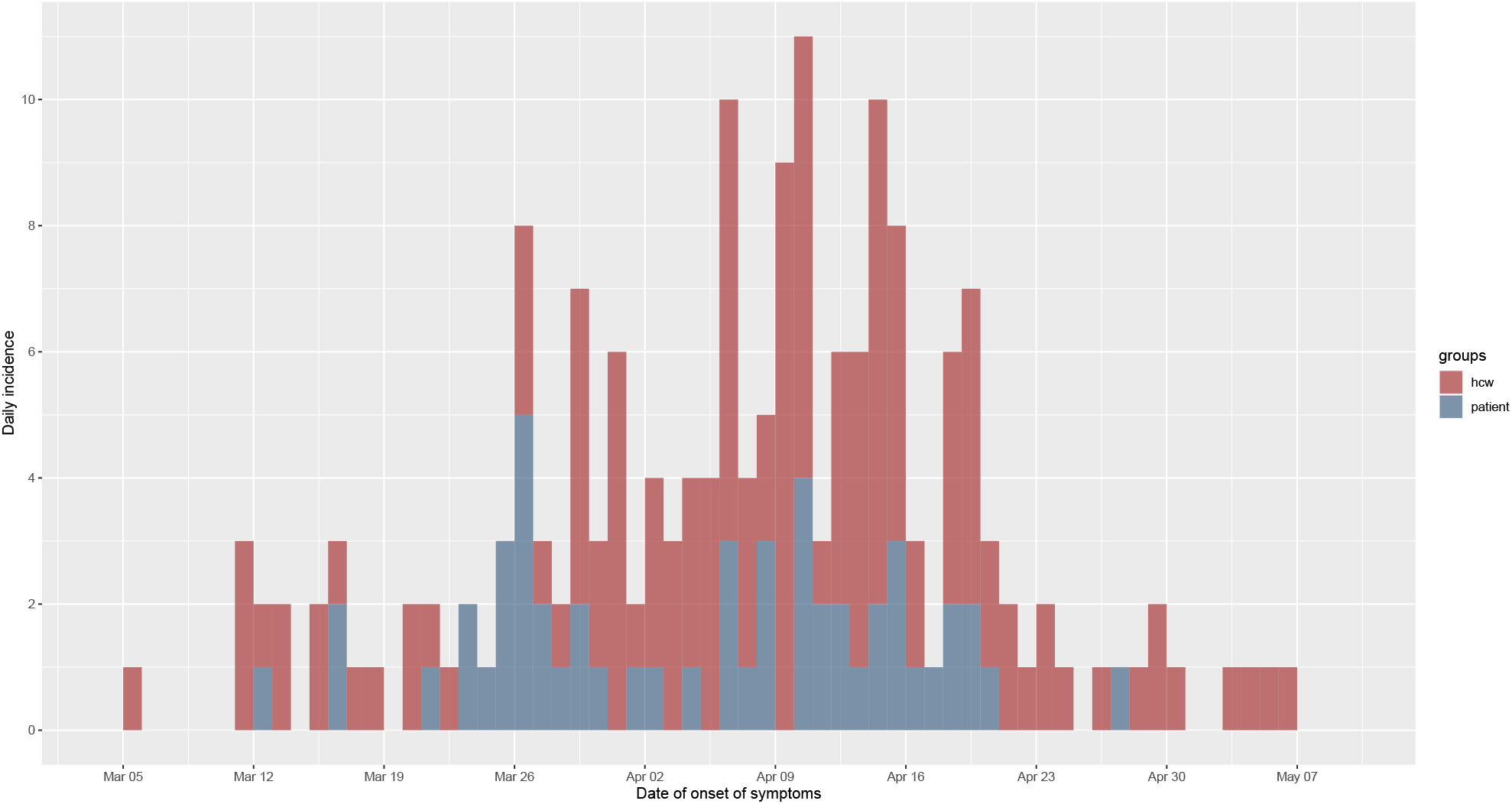
Epidemic curve of the nosocomial COVID-19 outbreak in a geriatric hospital involving HCWs and patients.

### Phylogenetic tree

We obtained SARS-CoV-2 sequences for 148 of the 180 cases (82.2%), including 105 HCWs (82.7%) and 43 patients (81.1%). A rooted phylogenetic tree suggests a large amount of genetic diversity, with at least 9 clusters and sub-clusters (Figure 2). One cluster (with moderate bootstrap support [BS] 26%) comprises sequences from 17 HCWs, 3 patients, and 6 community isolates in multiple subclusters (e.g. BS 72% with signature mutation C5239T). Another cluster (BS 78% with signature mutations C28854T & A20268G) shows a HCW sequence (H1048) with high similarity with community isolates. A large cluster (BS 68% with signature mutations C8293T, T18488C, and T24739C) with several subclusters includes 19 HCWs, 9 patients, and 3 community cases; ward movements for the patients are shown in Supplementary Figure S3A. A well-defined cluster (BS 100%) shows isolates from patients and HCWs from the same ward.

**Figure 2.**
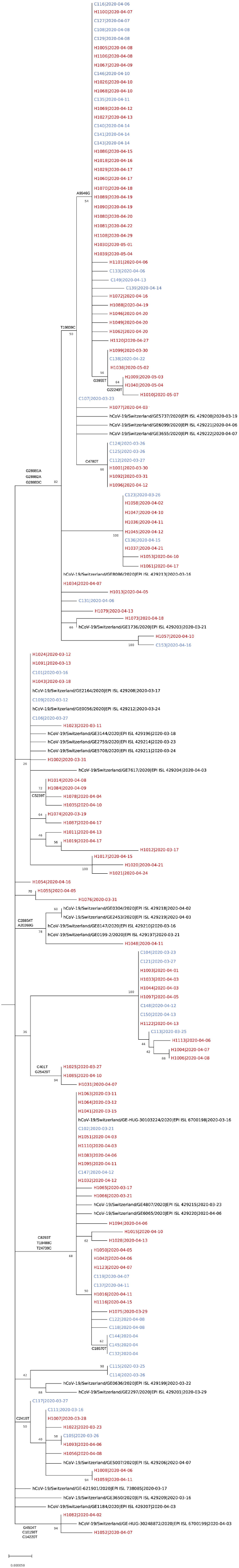
Phylogenetic tree of SARS-CoV-2 genome sequences. The tree includes 148 sequences related to the outbreak (patient and employee sequences are named C1xx [blue] and H10xx [red], respectively), alongside the community cases in the canton of Geneva, Switzerland, that were sequenced in March-April 2020 by the Laboratory of Virology (Geneva University Hospitals) and submitted to GISAID (virus names and accession ID (i.e. EPI_ISL_) are indicated) in the context of an epidemiological surveillance. For each sequence the date of the sample collection is mentioned (yyyy-mm-dd).

### Imported cases

In our reconstruction of who infected whom, we identified 16 imported cases of whom 13 were HCWs and three were patients (C107, C115, and C123, Table 3), two of which (C107 and C115) were initially classified as nosocomial. These 16 imported cases were found to have generated a total of 16 secondary cases, with 1 patient being involved in 5 onward transmissions. Together, imported cases and their direct secondary cases account for 21.6% of all cases.

**Table 3.** Imported cases and secondary infections

| Imported case | Probability of importation | Secondary onward transmission | Posterior probability of onward transmission |
| --- | --- | --- | --- |
| C115 | 0.60 | C114 | 0.60 |
| C107 | 1.00 | C131 | 0.70 |
|  |  | C124 | 0.39 |
|  |  | H1068 | 0.16 |
|  |  | H1005 | 0.33 |
|  |  | H1077 | 1.00 |
| C123 | 0.97 | H1058 | 0.89 |
| H1025 | 0.87 | H1085 | 0.94 |
| H1082 | 0.83 | H1052 | 0.83 |
| H1110 | 0.86 | H1064 | 0.32 |
|  |  | H1063 | 0.60 |
| H1008 | 0.84 | H1059 | 0.84 |
| H1073 | 1.00 |  |  |
| H1015 | 1.00 |  |  |
| H1011 | 0.66 | H1019 | 0.62 |
| H1012 | 1.00 |  |  |
| H1013 | 1.00 |  |  |
| H1122 | 0.86 | C113 | 0.57 |
| H1057 | 0.50 | C153 | 0.50 |
| H1048 | 1.00 |  |  |
| H1021 | 1.00 | H1017 | 1.00 |

### Reconstructing who infected whom

The output from our ancestry reconstruction is shown in Figure 3 (see Supplement for more results and sensitivity analyses). In addition to the 16 cases identified as imported, we were able to reconstruct with high confidence (>50% posterior probability) the ancestry of 36/132 (27.2%) of the remaining cases. The model estimated that the reporting probability was 91.5% (95% credible interval [CrI] 91.2%-91.9%), suggesting that only 8.5% of cases involved in transmission were unidentified. For most (92.0%) cases, the model identified the direct infector, without intermediate unobserved cases.

**Figure 3.**
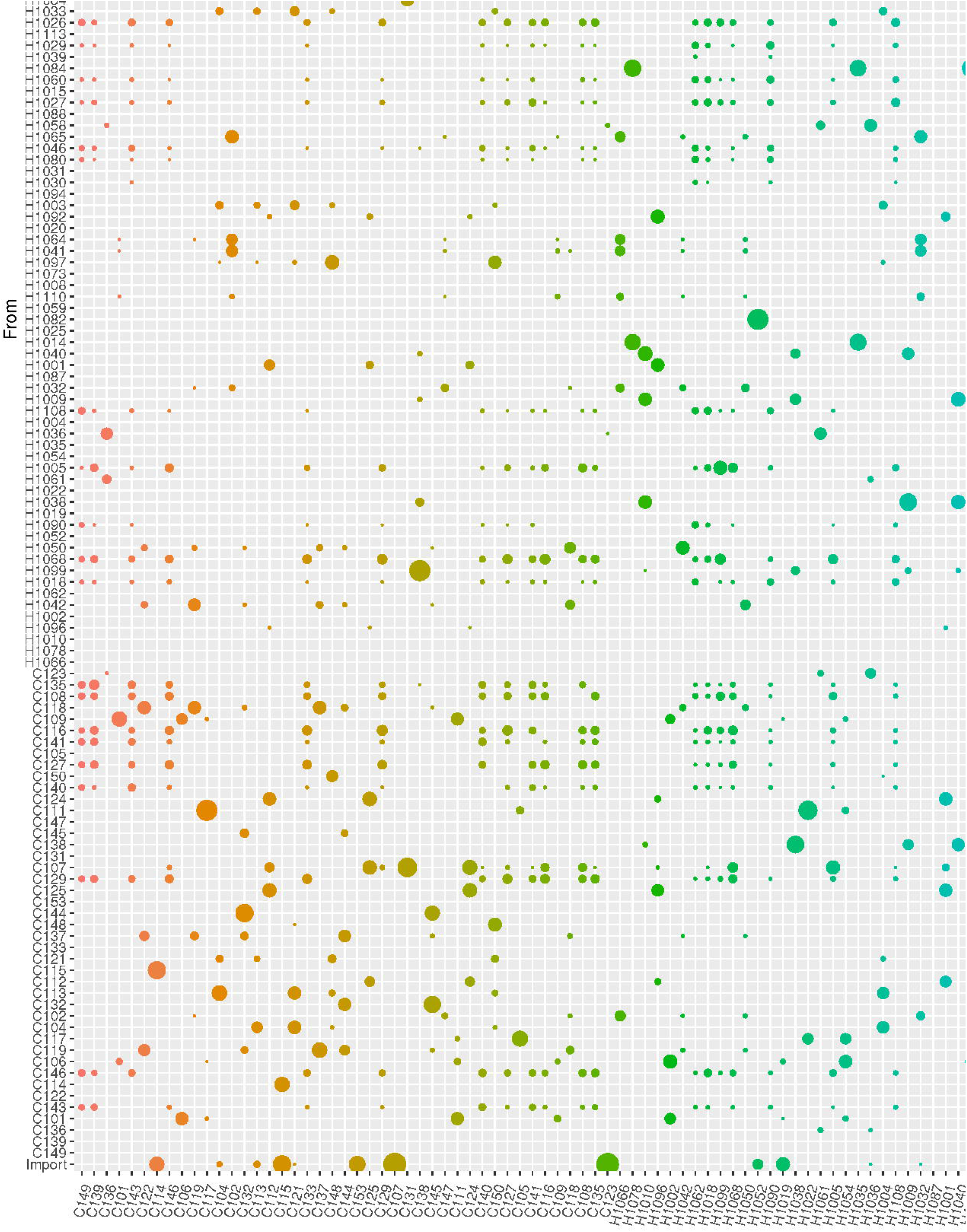
Ancestry reconstruction (who infected whom) of the `outbreaker2` model. Infectors are on the vertical axis and infectees are on the horizontal axis. Each bubble represents the posterior probability of each infector-infectee transmission pair. The bottom row denotes the probability that an infectee was in fact an imported case. Patients and employees are named C1xx and H10xx, respectively.

### Ward attribution

There was 95% agreement between the epidemiological attribution of the presumptive ward of infection (for patients) and that suggested by the model output (see Supplement). For 3 patients, the ward attribution was modified by the modelling analysis.

### Transmission patterns

Among patient-to-patient transmission events, and across all posterior trees, 97.6% (95%CrI 90.4-100%) involved patients who had shared a ward during their hospital stay. In contrast, only 44.4% (95%CrI 27.8-62.5%) of patient-to-patient transmissions involved patients who had shared a room. The model predicted that C107 infected C131 with a 70.5% probability although they did not share a ward (Supplementary Figure S3B); the probabilities that this was a direct infection and indirect infection with an unreported intermediate infector were 36.2% and 34.3%, respectively (Supplementary Figure S3B).

### Secondary infections

The number of secondary infections caused by each infected case (individual reproductive number, estimated from the transmission tree reconstruction), ranged from zero to nine (Figure 4). We compared the proportion of cases with no secondary transmissions (“non-transmitters”) and of cases with ≥2 secondary transmissions (“high transmitters”) across types of cases and phase of the outbreak. We found that the proportion of non-transmitters among both HCW_outbreak_ and patient_noso_ was smaller in the early than in the late stage (approximately 30% in early and 50% in late phase for both groups), suggesting that the contribution of these groups to ongoing transmission decreased over the study period. Conversely, the proportion of non-transmitters among HCW_covid_ was stable at about 55% across the early and the late phase. The proportions of high transmitters were significantly higher among HCW_outbreak_ than either patient_noso_ or HCW_covid_ in the late phases (26.2% vs. 13.4% and 11.4%, p<2.2e-16) of the outbreak. These trends were similar in the sensitivity analyses (Supplementary Table S2).

**Figure 4.**
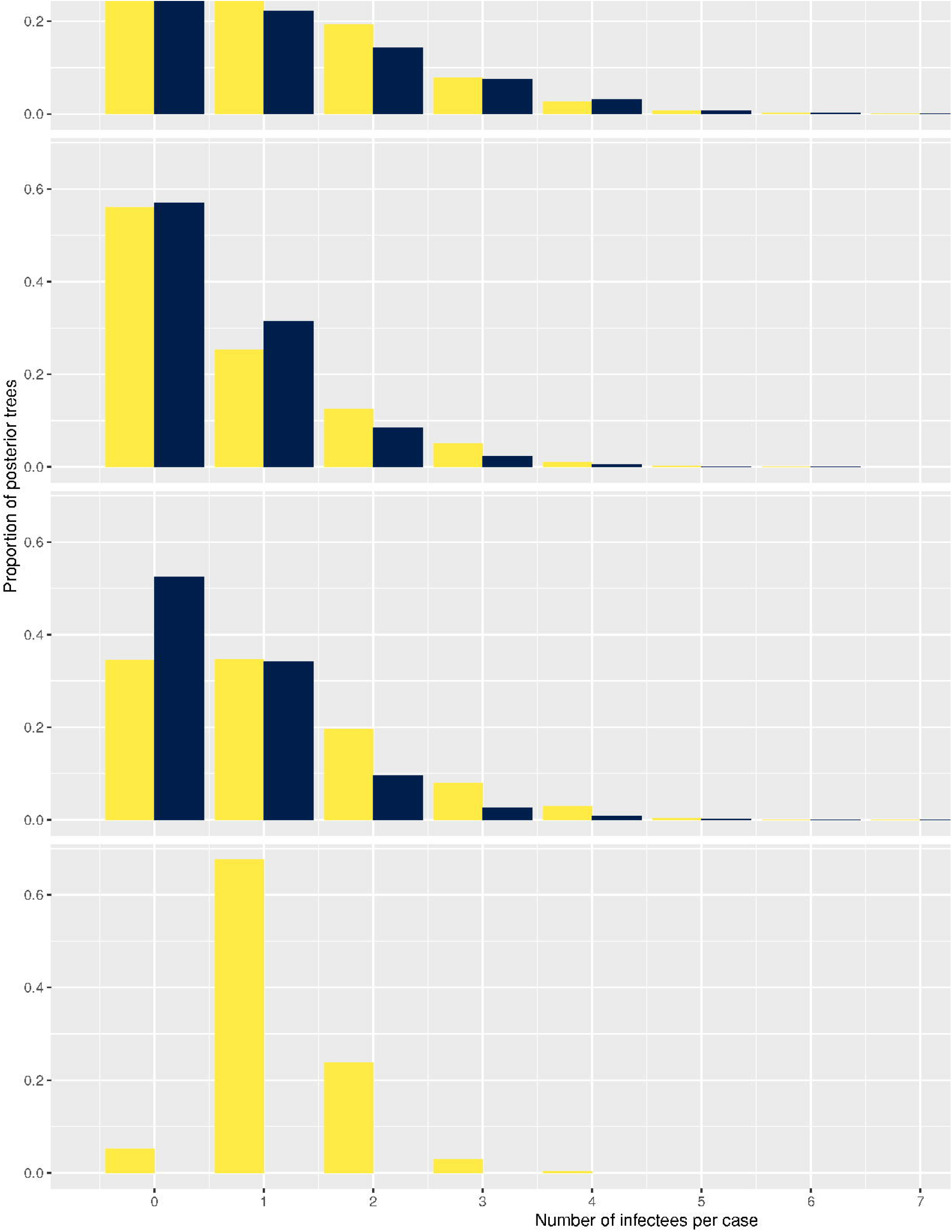
Histograms of distribution of secondary cases by each case type (“HCW_covid_”, HCWs working in Covid-19 wards; “HCW_outbreak_”, HCWs working in outbreak wards; “patient_noso_”, patients with hospital-acquired Covid-19; “patient_community_”, patients with community-acquired Covid-19) and stratified according to early (up to April 09, 2020) and late phases (as of April 10, 2020). Number of cases in early phase: HCW_outbreak_ 19, HCW_covid_ 43, patient_noso_ 25, patient_community_ 1. Number of cases in late phase: HCW_outbreak_ 7, HCW_covid_ 36, patient_noso_ 17, patient_community_ 0.

### Role of HCWs and patients in transmission events

We found that cases were significantly less likely than expected at random to be infected by HCWs from COVID wards (proportion infected by HCW_covid_, *f*_HCW_ = (43%; 95%CrI 36-49% versus 53% expected at random; 95%CrI 44-62%; p = 0.043). This was true across all cases, but particularly among HCWs in outbreak wards (*f*_HCW_ = 31%, 95% CrI 16-46%; p = 0.07) and patients, (*f*_*HCW*_ = 26%, 95%CrI 14-37%, p = 0.005). Conversely HCW_outbreak_ were significantly more likely than expected at random to become infected by other HCW_outbreak_ (proportion infected by HCW_outbreak_, *f*_outbreak_ = 38%, 95%CrI 24-54% versus 18%; 95%CrI 4-35%, p = 0.03). Patients with nosocomial Covid-19 (patient_noso_) were significantly more likely than expected at random to be infected by other patient_noso_ (proportion infected by patient_noso_, *f*_pat_ = 54%, 95%CrI 40-69% versus 28%; 95%CrI 14-45%, p = 0.01). Full results are shown in Figure 5.

**Figure 5.**
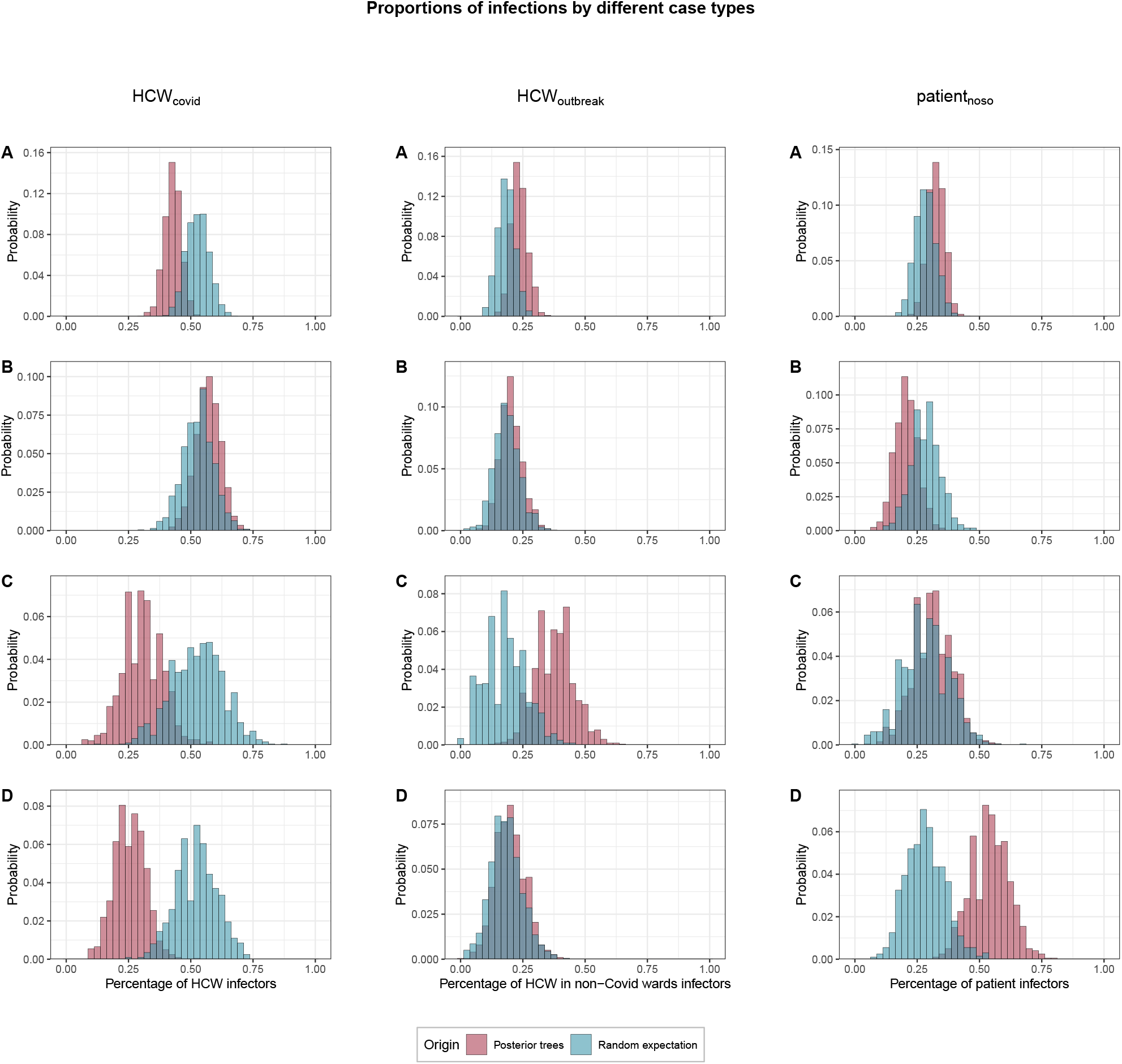
Proportions of transmissions (*f*_*case*_) attributed to each case type (HCW_covid_, HCW_outbreak_, patient_noso_, patient_community_) proportion of infections attributed to the type of case for each of the 1000 posterior trees retained. The blue histograms indicate the expected random distributions of *f*_*ward*,_ given the prevalence of each type of case. The red histograms show the observed distribution of *f*_*ward*,_ across 1000 transmission trees reconstructed by `outbreaker2`. **A**. All cases. **B**. Transmission to HCWs in Covid-19 wards only. **C**. Transmission to HCWs in non-Covid-19 wards (i.e. outbreak wards) only. **D**. Transmission to patients with nosocomial Covid-19 only.

### Role of within-ward and between-ward transmission

Infected staff or patients in outbreak wards were responsible for significantly more transmission in general (54%; 95%CrI 48-61%) than expected (43.7%, 95%CrI 35-53%). This was driven in particular by transmission from staff or patients to other staff or patients in outbreak wards (73%, 95%CrI 63-82%) (Figure 5). Within-ward transmission was more pronounced in outbreak wards (mean 48%; range: 20-70% of all infections within a ward) than in non-outbreak wards (mean 14%; range 0-63%) as shown in Supplementary Figures S4 and S5.

## Discussion

This in-depth investigation of SARS-CoV-2 transmission between patients and HCWs in a geriatric hospital, including several nosocomial outbreaks, has provided many valuable insights on transmission dynamics. First, we showed that the combination of epidemiological and genetic data using sophisticated modelling was able to tease out overall transmission patterns. Second, we showed that transmission dynamics among HCWs differed according to whether they worked in Covid wards or in wards where outbreaks occurred: while there was no excess HCW-to-HCW transmission in Covid wards, we observed a two-fold higher risk of transmission between HCWs in non-Covid wards. Third, we identified excess patient-to-patient transmission events, most of which occurred within the same ward, but not necessarily the same room. Fourth, we identified multiple importation events mainly from HCWs, but also from one patient with community-acquired Covid-19, that led to a substantial number of secondary cases and/or clusters.

These results are particularly important, as settings which care for elderly patients, such as geriatrics and rehabilitation clinics or LTCFs have high attack rates of SARS-CoV-2 for both patients and HCWs [7]. In an institution-wide seroprevalence study in our hospital consortium, the Department of Rehabilitation and Geriatrics, of which the hospital in this study was part of, had the highest proportion of HCWs with anti-SARS-CoV-2 antibodies [20].

The different transmission patterns between HCWs in Covid wards and outbreak wards (i.e. meant to be “Covid-free”) is intriguing. One hypothesis may be behavioural; indeed, it has been shown that HCWs caring for Covid-19 patients had significant concerns about being infected by their patients, and therefore may apply IPC measures more rigorously than when not caring for Covid-19 patients [21]. It may be that HCWs working in non-Covid wards did not feel threatened by Covid-19 patients on other wards, and thus not in their direct care. Also, it may be that HCWs underestimated the transmission risk from their peers to a greater extent in non-Covid wards than in Covid wards, leading to less physical distancing. Another contributing factor, which gives credence to the abovementioned hypotheses, is the higher mean duration of presenteeism despite symptoms compatible with Covid-19 in HCWs in non-Covid wards (2.9 days) compared to HCWs in Covid wards (1.6 days). There may be other factors (e.g. work culture, baseline IPC practices) that contribute to explain this difference in transmission patterns.

Most patient-to-patient transmission events involved patients having shared a ward, and were therefore in close proximity. Although we cannot exclude transmission from a “point-source” or hand borne transmission (i.e. an unidentified HCW infecting multiple patients in the same ward), there is little evidence to suggest that this is the case; indeed, these transmission patterns were robust to changes in the model assumptions. Mathematical models suggest that single-room isolation of suspected cases could potentially reduce the incidence of nosocomial SARS-CoV-2 transmission by up to 35% [22]. In the outbreaks we describe, symptomatic patients were identified promptly, with a median delay of 0 days between symptom onset and first positive swab. This may, however, not be sufficient as patients may be able to transmit pre-symptomatically to other patients [23]; thus, forecasting patients at high risk of developing nosocomial Covid-19 [24] may be useful should single-rooms not be available for all contacts of positive cases (e.g. in cases of overcrowding). Indeed, it has been suggested that exposure to community-acquired cases (i.e. already identified and therefore segregated/cohorted) was associated with half the risk of infection when compared to exposure to hospital-acquired cases or HCWs (who may be asymptomatic) [25]. One possible explanation is that by the time CA-Covid cases are hospitalised, they have passed the peak of infectiousness, whereas HA-Covid cases are in frequently unprotected contact with HCWs and other patients during their period of peak infectiousness.

It is our view that there is currently no strong evidence for the use of real-time genomics for control of SARS-CoV-2 nosocomial outbreaks [26]. Nevertheless, in this investigation, as in many others, we performed WGS to investigate transmission patterns [12]. Although we were able to gain considerable insight from the powerful combination of genetic sequencing data and rich epidemiological data, the outbreaks were successfully controlled without the availability of WGS, as was the case in many published reports [27, 28]. Furthermore, WGS may be more useful for ruling out transmission rather than for confirmatory purposes, due to the low number of mutations accumulated in the SARS-CoV-2 genome between transmission pairs [29].

Our study has several strengths. To the best of our knowledge, it is the only extensive outbreak investigation with WGS and sophisticated modelling in a geriatric acute-care hospital. Our WGS coverage was very high with 80% of cases involved providing sequencing data, including sequences from HCWs which has been previously shown to improve understanding of transmission dynamics [8]. The data collected for the purposes of this study were prospectively collected, minimising the risk of bias.

Despite these strengths, some limitations must be acknowledged. First, we did not include sequences from CA-Covid cases, bar one. However the method we used to reconstruct who infected whom is able to cope with and identify missing intermediate cases; here we estimated that the overwhelming majority of cases (91.5%) was captured in our sample. Another limitation is that these investigations were performed during the first pandemic wave in a susceptible population, and therefore the results may no longer be applicable in settings with high vaccination coverage and/or substantial natural immunity. Nevertheless, the lessons learned may be useful in a large number of countries with slow vaccine roll-out due to vaccine hesitancy, particularly in HCWs where there is no vaccine mandate, or unequal access to vaccine supplies [30]. Furthermore, nosocomial outbreaks of SARS-CoV-2 still occur despite high vaccination coverage [31, 32]. Also, these valuable lessons may be applicable for nosocomial outbreak control in the case of future pandemics due to respiratory viruses with characteristics similar to SARS-CoV-2.

In conclusion, strategies to prevent nosocomial SARS-CoV-2 transmission in geriatric settings should take into account the complex interplay between HCWs in dedicated Covid-19 wards versus non-Covid wards, and the potential for patient-to-patient transmission.

## Data Availability

Due to small size of the various clusters, clinical data will not be shared in order to safeguard anonymity of patients and HCWs.

## Acknowledgements

We would like to thank Frédéric Bouillot (Geneva University Hospitals) for providing us with HR data, Aurore Britan (Geneva University Hospitals) for data management, Rachel Goldstein for data collection (Geneva University Hospitals), Daniel Teixeira for data extraction (Geneva University Hospitals). In addition, the authors would like to thank Stéphanie Baggio, Frédérique Jacquerioz Bausch, Hervé Spechbach from the AMBUCoV study (Geneva University Hospitals), Amaury Thiabaud (University of Geneva), as well as the IPC team members from Geneva University Hospitals involved in SARS-CoV-2 outbreak management (Pascale Herault, Didier Pittet, Walter Zingg). Ashleigh Myall (Imperial College London) provided code to help generate the edge list of contacts. Thibaut Jombart (London School of Hygiene and Tropical Medicine) and Finlay Campbell (World Health Organization) provided input on technical aspects of the `outbreaker2` model development. The study team also wishes to thank the dedicated HCWs who looked after patients professionally, as well as the Geriatric Hospital Covid-19 crisis management team (Gabriel Gold, Charline Couderc, Etienne Satin).

