## Supplementary for "Reconstructing transmission chains of SARS-CoV-2 amid multiple outbreaks in a geriatric acute-care hospital"

This appendix has been provided by the authors to give readers additional information about their work.

Table of contents

Supplementary Material 3

**Infection prevention and control measures during the 1st pandemic wave** **3**

**Microbiological Methods** **4**

*Amplicon-based high-throughput sequencing (HTS) analysis* 4

*Phylogenetic analysis* 4

**Statistical analyses** **5**

*Implementation of the outbreaker2 models* 5

Supplementary Tables 9

**Supplementary Table 1.** Composition of baseline `outbreaker2` model and different sensitivity analyses. **9**

**Supplementary Table 2.** Proportions of secondary infections (i.e. individual *R*) for each case type ($patient_{noso}$, $HCW_{outbreak}$, $HCW_{covid}$) in early (up to April 09, 2020) and late (as of April 10, 2020) phases of the study. The p-values are for chi-squared tests on these proportions. **10**

Supplementary Figures 12

**Supplementary Figure 1.** Ward-level epidemic curve. **12**

**Supplementary Figure 2.** Comparison of the accuracy of ancestry attribution of each sensitivity analysis. **13**

**Supplementary Figure 3.** Ward movements for patients involved in a cluster. **14**

**Supplementary Figure 4.** Ward-to-ward transmission matrix. **15**

**Supplementary Figure 5.** Proportions of transmissions attributed to outbreak ($f_{outbreak\text{-}ward}$) and non-outbreak ($f_{non\text{-}outbreak\text{-}ward}$) wards. **16**

References 17

Supplementary Material

### Infection prevention and control measures during the 1st pandemic wave

A multidisciplinary Covid-19 response unit was formed in the Geriatric hospital, with daily meetings and involvement of the IPC team. Direct coaching of HCWs working in Covid-19 wards by the IPC practitioner was undertaken once a ward had been attributed as such. The IPC practitioner assisted front-line HCWs in streamlining tasks.

HCWs caring for Covid-19 patients applied "contact" and "droplet" precautions, in line with Swissnoso and FOPH guidelines. Universal masking of all front-line HCWs was implemented on March 11, 2020. From April 01, 2020, the use of ocular protection (eye shields) was encouraged for contact with all Covid-19 patients, and masking of HCWs in non-clinical areas (e.g. offices) was recommended.

RT-PCR screening of SARS-CoV-2 on admission for all patients, even if asymptomatic or without clinical suspicion of Covid-19, started on April 01, 2020. From April 07, 2020, weekly screening surveys were performed in non-Covid wards, until May 30, 2020.

As of April 11, 2020 non-Covid wards were closed to new admissions, and room occupancy was decreased (e.g. 4-bed rooms were limited to 3 patients).

Due to shortages in PPE, HCWs were instructed to wear surgical masks for 4 hours continuously (and 8 hours if not humid), and N95 respirators (FFP2 masks) for as long as possible. Gowns were used for >1 patient in the same room, except in the case of patients carrying multidrug resistant bacteria.

HCWs from outbreak wards were encouraged to undergo PCR testing on nasopharyngeal swabs, even if asymptomatic between April 09 and April 16, 2020. Including tests that were performed since mid-March, a total of 83 out of 124 eligible HCWs (67%) in the four outbreak wards underwent testing.

### Microbiological Methods

*Amplicon-based high-throughput sequencing (HTS) analysis*

All nasopharyngeal swabs (NPS) tested positive for SARS-CoV-2 by RT-PCR (Cobas 6800 SARS-CoV2 RT-PCR), the Charite or the BD SARS-CoV2 reagent kit for BD Max system assays, and for which sufficient volume remained, were selected for whole-genome sequencing analysis.

NPS were sequenced with an amplicon-based sequencing method. Thus, nucleic acids were extracted using the NucliSENS easyMAG (bioMérieux, Geneva, Switzerland) and then sequenced using an updated version of the nCoV-2019 sequencing protocol (https://www.protocols.io/view/ncov-2019-sequencing-protocol-bbmuik6w) (Microsynth, Balgach, Switzerland) on a MiSeq instrument (Illumina) with a 2x250-bp protocol.

Duplicate reads were removed using cd-hit (v4.6.8). Low-quality and adapter sequences were trimmed out using Trimmomatic (v0.33). Reads were then mapped against the reference sequence MN908947 using snap-aligner (v1.0beta.18). Consensus for sequences with at least 10-fold coverage were then generated using custom script.

*Phylogenetic analysis*

Sequence alignment was performed with MUSCLE (v3.8.31). Evolutionary analyses were conducted in MEGA X [1] using the Maximum Likelihood method and Tamura 3-parameter model [2]. The tree includes also all SARS-CoV-2 complete genomes sequenced by our laboratory and submitted to GISAID from respiratory samples from COVID-19 positive patients presenting to our institution or other medical centres in Geneva, Switzerland, during the same period.

### Statistical analyses

#### *Implementation of the outbreaker2 models*

We combined epidemiologic and genetic data using the outbreaker2 package in the R software, which has been used successfully in the reconstruction of the 2003 SARS-CoV-1 outbreak in Singapore [3, 4] and a nosocomial outbreak of SARS-CoV-2 in a rehabilitation clinic [5]. The model uses a Bayesian framework, which combines information on the generation time (time between infections in an infector/infectee pair), with a model of sequence evolution to probabilistically reconstruct the transmission tree.

As dates of onset of symptoms are known, and because dates of infection (i.e. acquisition) are not known with certainty, we imputed serial intervals based on estimates from the work by Ali et al. [6], who showed that the serial interval decreased from the early stages of the pandemic due to improved control using non-pharmaceutical measures. For the primary analysis we used a short serial interval (mean 3.0, standard deviation [SD] 4.1), under the assumption of swift isolation of patients following onset of symptoms. We also performed a sensitivity analysis using a longer serial interval (mean 5.2 days, SD 4.7) to allow for potentially slower isolation of symptomatic patients. We used the incubation period as estimated by Bi et al., which follows a lognormal distribution with parameters mu of 1.57 and sigma 0.65 (corresponding to a mean of 5.95 days and SD 4.31) [7]. Where dates of onset were unavailable, we imputed them by using the median of the difference between symptom onset and date of swab.

Imported cases are detected by outbreaker2 during an initial step of the model analysing the global influence of each case based on its genetic log-likelihood [3]. By default, a case is determined as being imported if its global influence is 5 times the average global influence. While this approach has excellent specificity, because of the limited genetic diversity of SARS-CoV-2, it may lack in sensitivity. We therefore ran the model over a range of lower thresholds (from 2 to 5 times the average global influence). We selected for our main model a threshold of 2, and present one-way sensitivity analyses with a threshold of 3 and the default (5).

The outbreaker2 package is designed to use contact tracing data to inform who infected whom. These data were unavailable for our outbreak, but we made a series of assumptions to generate a matrix of possible contacts between cases. Initially, we aimed to construct this matrix using dates of presence in the hospital/ward for patients based on administrative data, and based on human resources shift rota for HCWs. However, we identified potential inconsistencies in the latter, in particular stemming from multiple changes as a result of many HCWs self-isolating due to possible or confirmed COVID-19. We therefore used these data in a sensitivity analysis, but for our main analysis, we reverted to a simpler set of assumptions to build our contact matrix. In our main analysis, we assumed that HCWs were present in the hospital every day until the date of their first positive swab, included. We further assumed that patients only interacted with patients in their own ward. We assumed that all HCWs were able to infect each other, but HCWs could have contact with patients only in the wards they were attributed to; HCWs such as physical therapists or doctors who worked across multiple wards could have significant contact with all HCWs and patients. Under these assumptions, we are likely to capture many contacts which did not happen, and it is possible that we miss a few contacts which in fact did happen. However outbreaker does account for imperfect sensitivity and specificity of contact data, the levels of which are estimated as part of the model. To account for this uncertainty in potential contact patterns, we ran 2 sensitivity analyses where we input different contact data in the model.

#### Sensitivity analyses

We conducted a number of different analyses, including a base scenario and several (n = 5) one-way sensitivity analyses (Supplementary Table 1):

- Sensitivity analysis 1:
    - We did not make any assumptions about contact patterns, and therefore all cases in the outbreak could potentially infect all other cases.
- Sensitivity analysis 2:
    - We used a longer serial interval (mean 5.2 days, SD 4.7) to allow for potentially slower isolation of symptomatic patients.
- Sensitivity analysis 3:

- We used the HCW shift data from the human resources department, with minor corrections (removing HCWs that were mislabelled as "present" after date of positive RT-PCR). For both patients and HCWs we categorised days of "susceptibility" (5th percentile of the cumulative incubation period from Bi et al. [7]) and days of "infectiousness" (2 days before symptom onset based on the study by He et al. [8]). The last day of "infectiousness" was the date of swab for HCWs.

- Sensitivity analysis 4:
  - We assumed that isolation precautions prescribed for patients on date of positive RT-PCR were effective, and that from that date patients were no longer infectious.
- Sensitivity analysis 5:
  - We used a higher threshold of 3 for the determination of imported cases.
- Sensitivity analysis 6:
  - We used the default threshold (5) used by outbreaker2 to detect imported cases.

For each model, we used a uniform prior between 0.55 and 1 for the reporting probability ("pi"). Indeed, we had a comprehensive screening and testing strategy, including of asymptomatic cases, and are therefore confident that we captured a near-total proportion of cases. We obtained sequences for 82% of all identified cases, and these are the cases used in the model. The lower bound of the prior for "pi" thus allows us to have missed 6 cases in addition to the 14 that were not sequenced. Posterior estimates for "pi" were visually compared to our prior choice to assess the validity of this assumption.

We allowed for a maximum of 2 unobserved cases on a transmission chain between any two observed cases (maximum "kappa" of 3 in outbreaker2). This allows for identification of missed cases.

We used the default priors for the mutation rate ("mu") for all models (uninformative exponential prior with mean 1), and, where relevant, those for non-infectious contact rate ("lambda") and contact reporting coverage ("eps"), which were uniform on [0, 1] [4, 9]. We used the default likelihoods for all models, except for the model without contact data where this was disabled [4, 9].

We ran each outbreaker model over 2,000,000 iterations of the MCMC (500,000 for model without contact data), with a thinning of 1 in 2000 (1 in 500 for model without contact data), in order to obtain a sample of 1000 posterior parameter sets, after a burn-in of 2000 iterations (500 for model with contact data). Each of these parameter sets corresponds to a posterior transmission tree. Convergence was assessed visually as well as through the Gelman-Rubin convergence diagnostic (using the gelman.diag function in the R package coda v0.19-4) [10], concluding that the chains converged appropriately if the upper limit of the confidence interval was < 1.1.

To assess the role of each type of case in transmission by estimating the proportion of infections attributed to the type of case ($f_{case}$), we compared the posterior distribution of direct infections caused by each infector type to that obtained by a matching number of random draws of the infector types (expected proportion of infections), drawn according to the prevalence of each type among cases. We concluded that $f_{case}$ was significantly higher than expected by chance if at least 95% of the posterior samples had higher proportions of cases infected by an infector type than that obtained by the random draws. The corresponding p-values were calculated as 1 minus the proportion of posterior samples with values higher than random draws. This was done for all infectees (i.e. whole outbreak), as well as for each type of infectee.

We evaluated the differences in the distribution of secondary cases across case types in the early (up to April 09, 2020) and later phases (as of April 10, 2020) of the study period using a chi-squared test (for proportions of cases with no secondary transmissions ("non-transmitters”) and of cases with ≥2 secondary transmissions ("high transmitters”)).

Supplementary Tables

**Supplementary Table 1.** Composition of baseline `outbreaker2` model and different sensitivity analyses.

| **Scenario type** | **Onset of symptoms** | **Genetic data** | **Contact data** | **Short serial interval** | **Longer serial interval** | **Low outlier threshold** | **Medium outlier threshold** | **Default outlier threshold** |
|---|---|---|---|---|---|---|---|---|
| Baseline scenario | X | X | X | X | | X | | |
| Sensitivity analyses | | | | | | | | |
| 1. | X | X | | X | | X | | |
| 2. | X | X | X | | X | X | | |
| 3. | X | X | X[a] | X | | X | | |
| 4. | X | X | X[b] | X | | X | | |
| 5. | X | X | X | X | | | X | |
| 6. | X | X | X | X | | | | X |

[a] For this model, we used the HR data for healthcare worker presence (with some corrections)

[b] For this model, we assumed that patients were no longer infectious after the date of positive RT-PCR

Supplementary Table 2. Proportions of secondary infections (i.e. individual *R*) for each case type ($patient_{noso}$, $HCW_{outbreak}$, $HCW_{covid}$) in early (up to April 09, 2020) and late (as of April 10, 2020) phases of the study. The p-values are for chi-squared tests on these proportions.

| | $patient_{noso}$ | $HCW_{outbreak}$ | $HCW_{covid}$ | p-value (overall) | p-value ($HCW_{outbreak}$ vs. $patient_{noso}$) |
|---|---|---|---|---|---|
| Main analysis | | | | | |
| ≥1 secondary transmission | | | | | |
| Early phase | 0.654 | 0.667 | 0.44 | <2.2e-16 | 0.006 |
| Late phase | 0.476 | 0.484 | 0.429 | <2.2e-16 | 0.26 |
| ≥2 secondary transmissions | | | | | |
| Early phase | 0.308 | 0.308 | 0.187 | <2.2e-16 | 0.96 |
| Late phase | 0.134 | 0.262 | 0.114 | <2.2e-16 | <2.2e-16 |
| Sensitivity analysis #1 (no assumptions about contacts) | | | | | |
| ≥1 secondary transmission | | | | | |
| Early phase | 0.692 | 0.659 | 0.443 | <2.2e-16 | 1.36E-13 |
| Late phase | 0.538 | 0.459 | 0.411 | <2.2e-16 | <2.2e-16 |
| ≥2 secondary transmissions | | | | | |
| Early phase | 0.367 | 0.283 | 0.181 | <2.2e-16 | <2.2e-16 |
| Late phase | 0.192 | 0.21 | 0.102 | <2.2e-16 | 0.001 |
| Sensitivity analysis #2 (long serial interval) | | | | | |
| ≥1 secondary transmission | | | | | |
| Early phase | 0.629 | 0.686 | 0.458 | <2.2e-16 | <2.2e-16 |
| Late phase | 0.469 | 0.477 | 0.385 | <2.2e-16 | 0.29 |
| ≥2 secondary transmissions | | | | | |
| Early phase | 0.346 | 0.342 | 0.206 | <2.2e-16 | 0.38 |
| Late phase | 0.149 | 0.265 | 0.131 | <2.2e-16 | <2.2e-16 |
| Sensitivity analysis #3 (calibrating contacts based on assumptions on infectiousness) | | | | | |
| ≥1 secondary transmission | | | | | |
| Early phase | 0.689 | 0.671 | 0.455 | <2.2e-16 | 7.28E-05 |
| Late phase | 0.455 | 0.352 | 0.328 | <2.2e-16 | <2.2e-16 |
| ≥2 secondary transmissions | | | | | |
| Early phase | 0.382 | 0.364 | 0.196 | <2.2e-16 | <0.001 |
| Late phase | 0.127 | 0.188 | 0.108 | <2.2e-16 | <2.2e-16 |
| Sensitivity analysis #4 (patients no longer infectious after date of swab) | | | | | |
| ≥1 secondary transmission | | | | | |
| Early phase | 0.61 | 0.672 | 0.442 | <2.2e-16 | <2.2e-16 |

| | $patient_{noso}$ | $HCW_{outbreak}$ | $HCW_{covid}$ | p-value (overall) | p-value ($HCW_{outbreak}$ vs. $patient_{noso}$) |
|---|---|---|---|---|---|
| Late phase | 0.48 | 0.485 | 0.397 | <2.2e-16 | 0.43 |
| ≥2 secondary transmissions | | | | | |
| Early phase | 0.311 | 0.311 | 0.186 | <2.2e-16 | 0.98 |
| Late phase | 0.143 | 0.259 | 0.129 | <2.2e-16 | <2.2e-16 |
| Sensitivity analysis #5 (higher value for outlier threshold) | | | | | |
| ≥1 secondary transmission | | | | | |
| Early phase | 0.618 | 0.666 | 0.435 | <2.2e-16 | <2.2e-16 |
| Late phase | 0.466 | 0.48 | 0.395 | <2.2e-16 | 0.053 |
| ≥2 secondary transmissions | | | | | |
| Early phase | 0.303 | 0.309 | 0.181 | <2.2e-16 | 0.21 |
| Late phase | 0.133 | 0.251 | 0.13 | <2.2e-16 | <2.2e-16 |
| Sensitivity analysis #6 (default value for outlier threshold) | | | | | |
| ≥1 secondary transmission | | | | | |
| Early phase | 0.657 | 0.679 | 0.445 | <2.2e-16 | 2.27E-06 |
| Late phase | 0.474 | 0.495 | 0.416 | <2.2e-16 | 0.005 |
| ≥2 secondary transmissions | | | | | |
| Early phase | 0.302 | 0.325 | 0.19 | <2.2e-16 | 2.92E-07 |
| Late phase | 0.136 | 0.265 | 0.157 | <2.2e-16 | <2.2e-16 |

Supplementary Figures

**Supplementary Figure 1.** Ward-level epidemic curve.

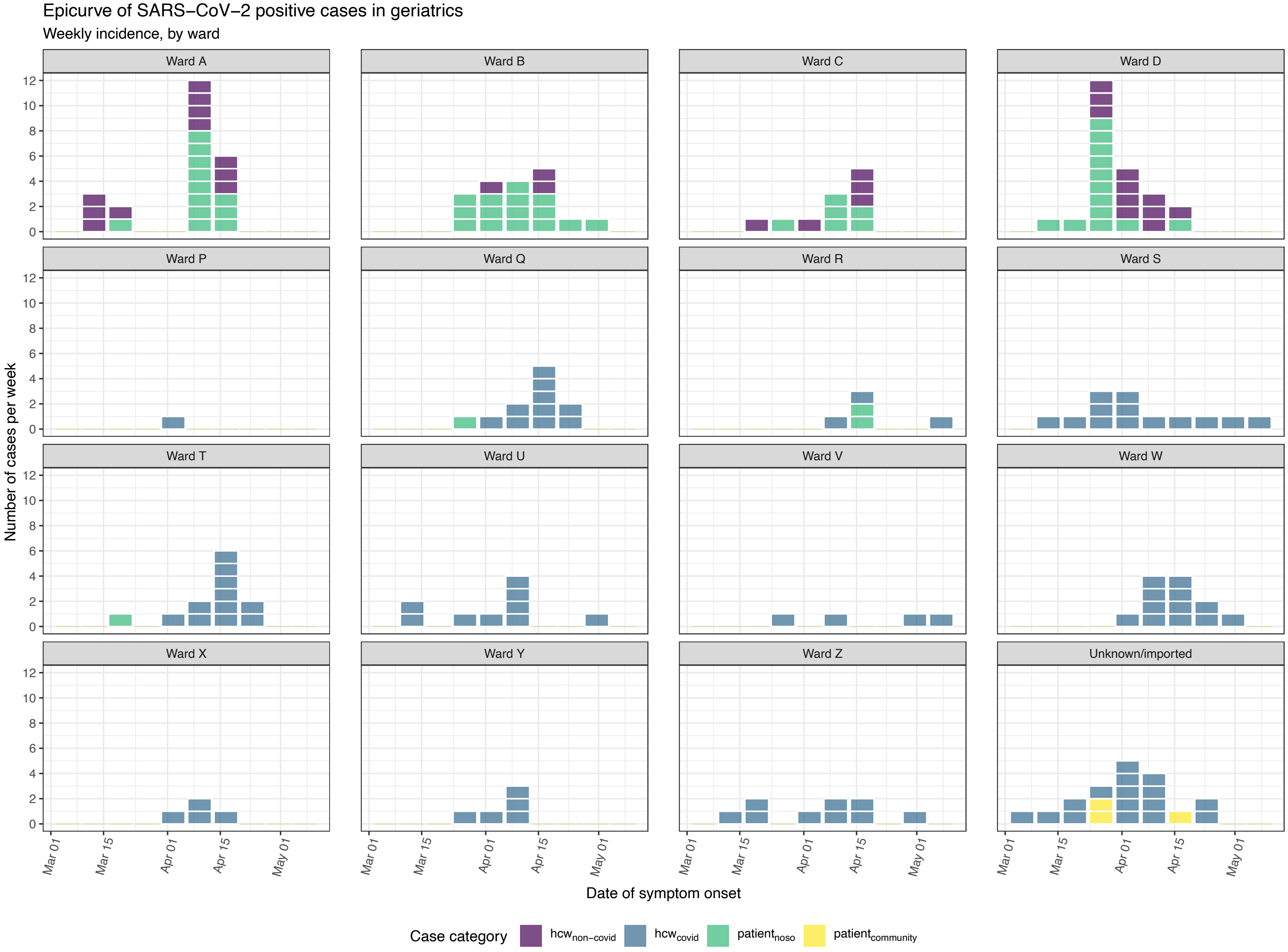

**Supplementary Figure 2.** Comparison of the accuracy of ancestry attribution of each sensitivity analysis.
Each case is on the horizontal axis. The shading corresponds to a value indicating the magnitude of the difference in attribution of the case's ancestor and the main analysis. For each infectee (case), we calculated the absolute difference in probabilities of infectors (ancestor) between the main analysis and each sensitivity analysis. Values of difference 1 indicate 100% difference in ancestry attribution, and 0 indicate absolute agreement.
Sensitivity analysis #1: absence of contact data.
Sensitivity analysis #2: longer serial interval (mean 5.2 days, SD 4.7) [7].
Sensitivity analysis #3: contacts were based on human resources data for HCWs and on infectious and susceptible periods.
Sensitivity analysis #4: patients are considered to be no longer infectious after the date of the positive RT-PCR for SARS-CoV-2.
Sensitivity analysis #5: higher value (3) for the threshold for identification of outliers.
Sensitivity analysis #6: default value (5) for the threshold for identification of outliers.

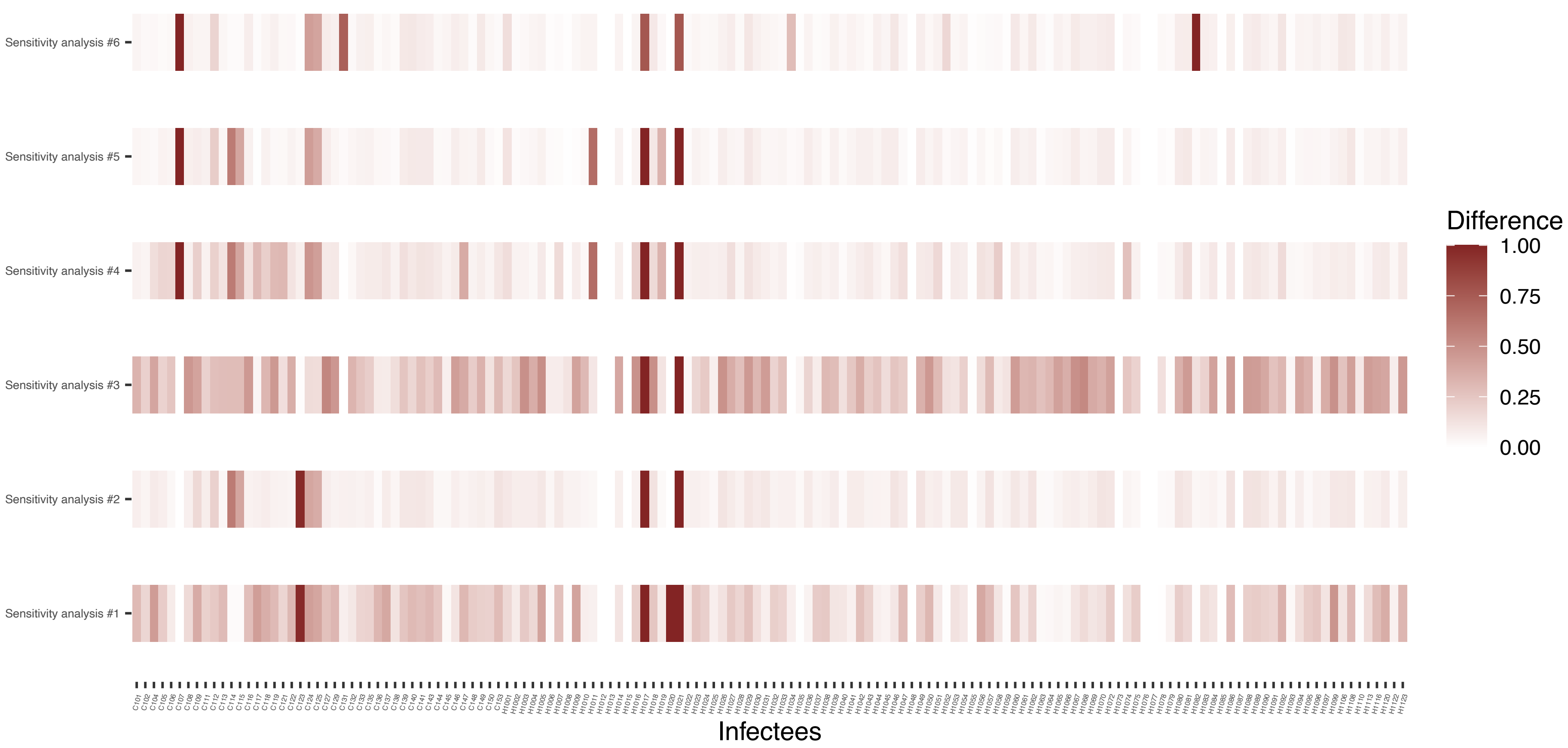

**Supplementary Figure 3.** Ward movements for patients involved in a cluster.
Each row corresponds to a patient, and the solid lines indicate hospitalisation dates. The lines are coloured according to which ward a patient was in on a particular day. Outbreak wards (A-D) are coloured differently from non-outbreak wards (Q-Z).

**A**

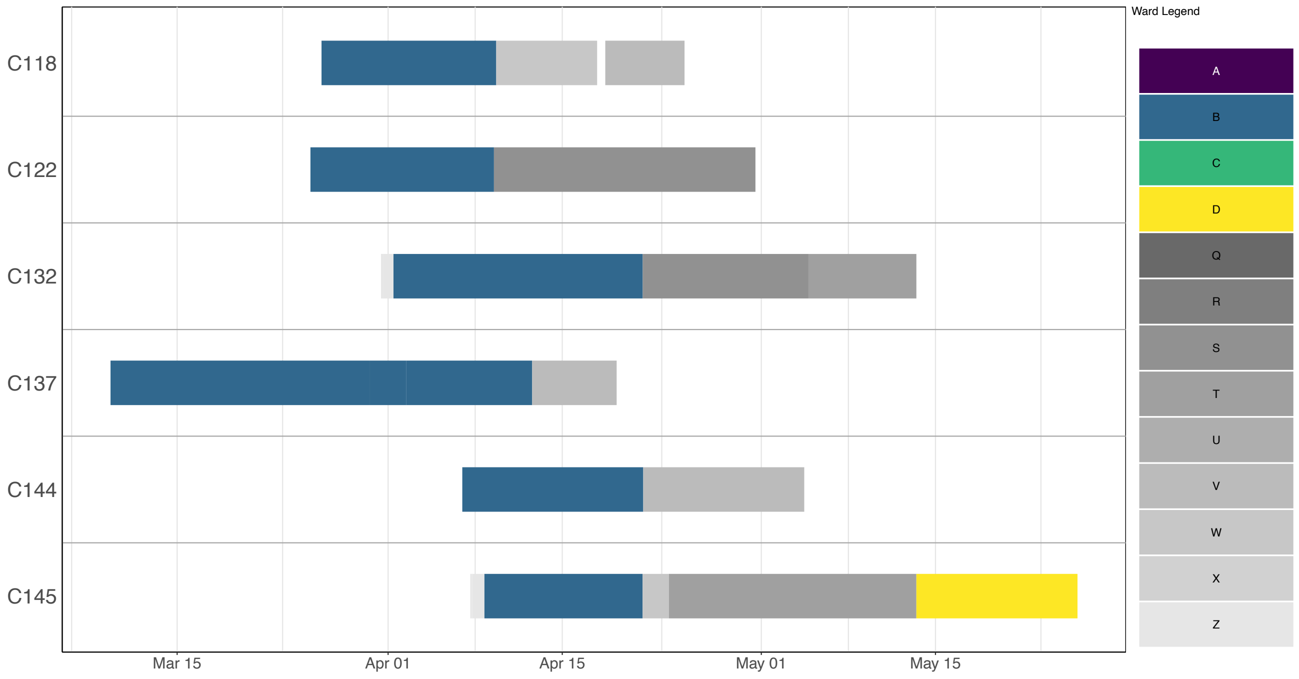

**B**

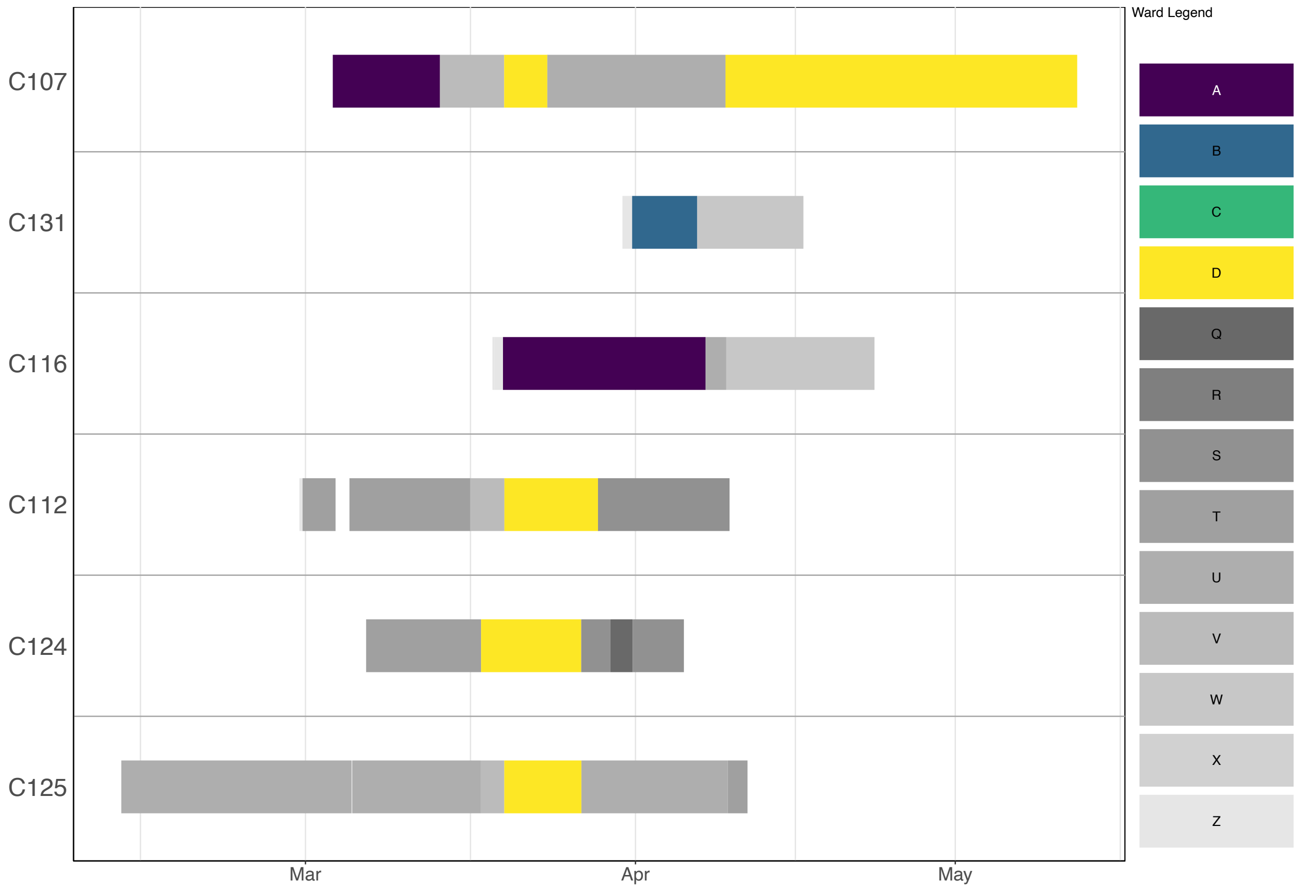

Supplementary Figure 4. Ward-to-ward transmission matrix.
The matrix indicates the sum of transmission events across all posterior trees from cases in "infector" wards (vertical axis) to cases in "infectee" wards (horizontal axis). The degree of shading is proportional to the estimated posterior number of transmissions for each ward-to-ward pair. Outbreak wards: A-D; non-outbreak wards: P-Z (Z is "all wards").

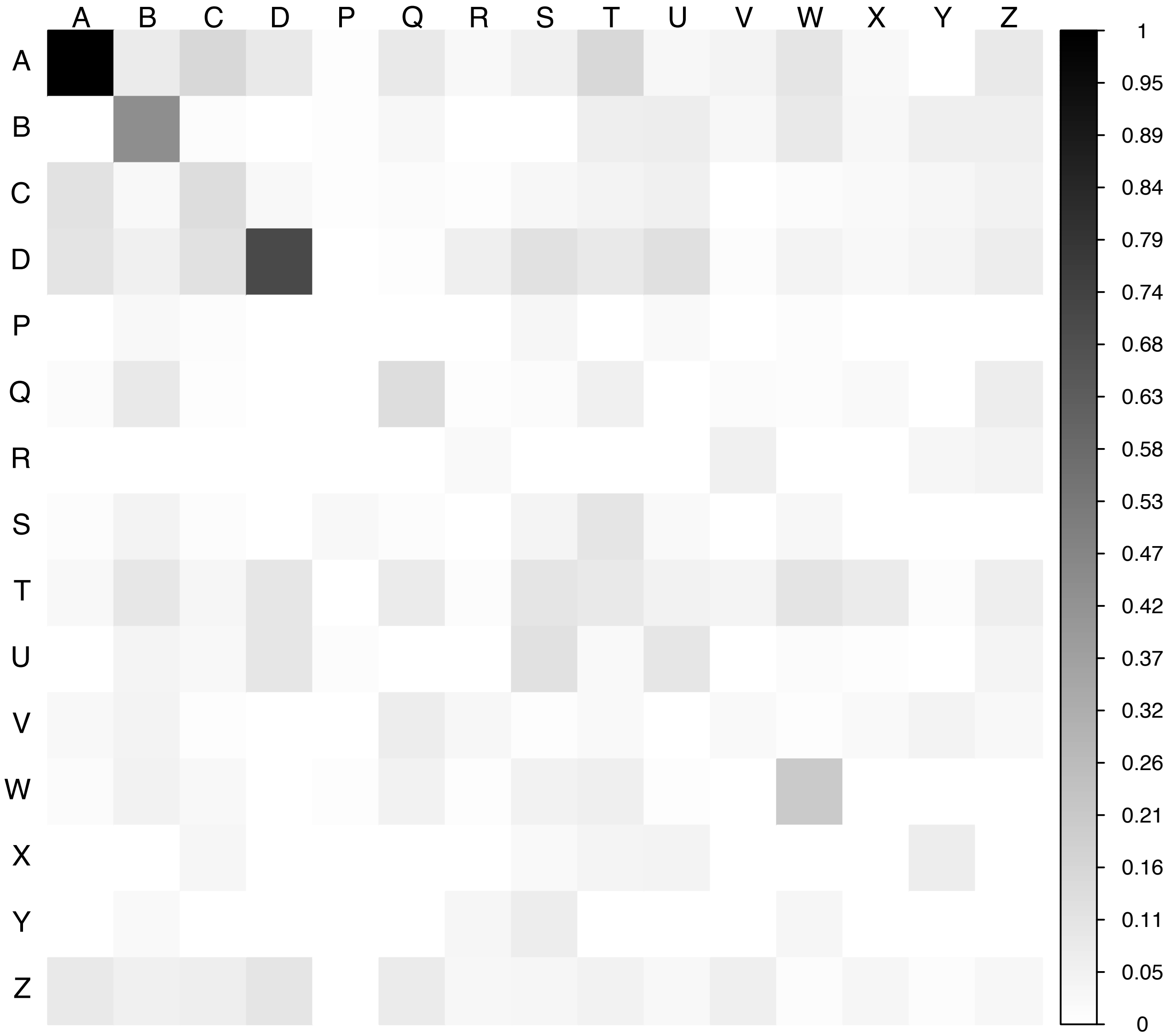

**Supplementary Figure 5.** Proportions of transmissions attributed to outbreak ($f_{outbreak\text{-}ward}$) and non-outbreak ($f_{non\text{-}outbreak\text{-}ward}$) wards.

The blue histograms indicate the expected random distributions of $f_{ward,}$ given the proportion of HCWs amongst cases. The red histograms show the observed distribution of $f_{ward,}$ across 1000 transmission trees reconstructed by `outbreaker2`. **A.** All wards. **B.** Transmission to outbreak wards only. **C.** Transmission to non-outbreak wards only.

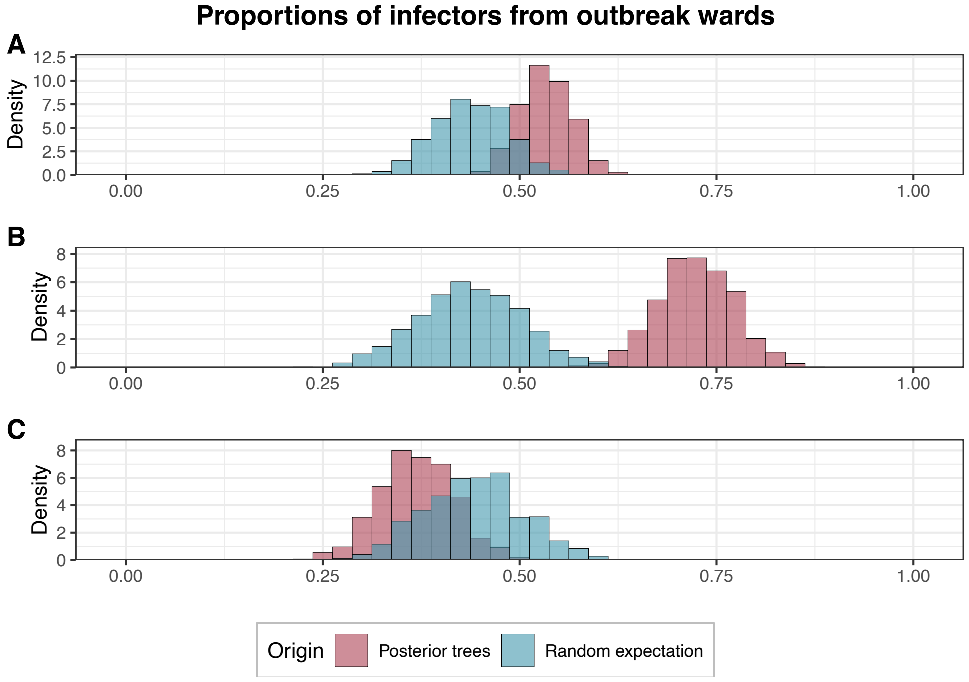

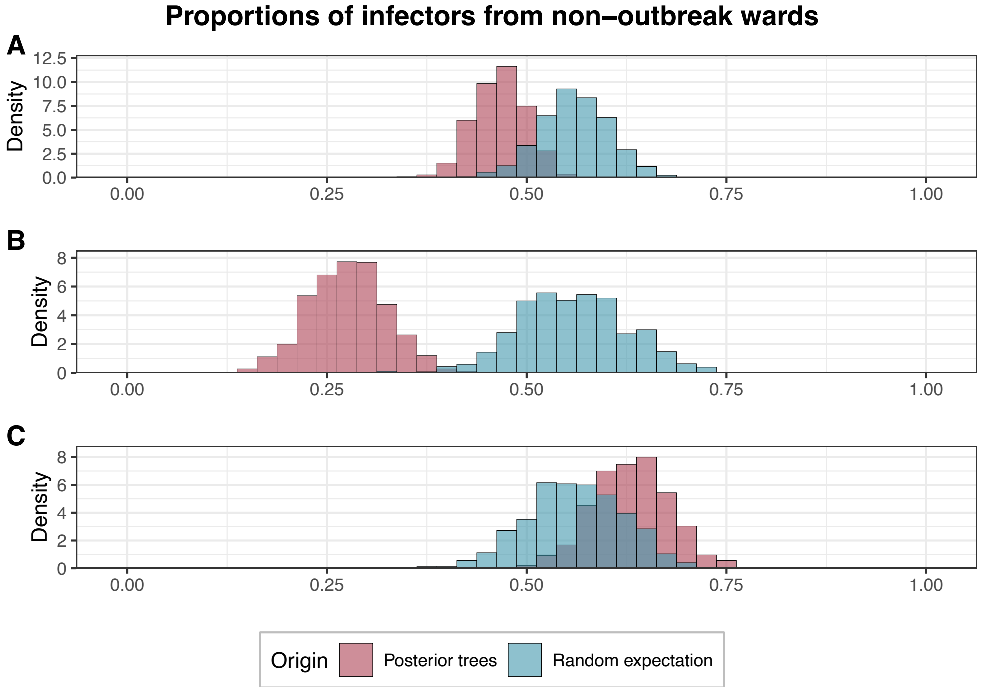
